# Exercise snacks are feasible to perform in the real world and improve physical capacity for adults living with non-insulin treated type 2 diabetes: a randomized trial

**DOI:** 10.64898/2026.02.23.26346877

**Authors:** Fiona J. Babir, Alexis Marcotte-Chénard, Roderick E. Sandilands, Kaja Falkenhain, Noah Mulkewich, Hashim Islam, Seth F. McCarthy, Douglas L. Richards, Kenneth Madden, Joel Singer, Michael C. Riddell, Mary E. Jung, Martin J. Gibala, Jonathan P. Little

## Abstract

**Aims/hypothesis:** To investigate the feasibility and preliminary efficacy of a 12-week remotely-delivered exercise snacks (ES) intervention in adults with type 2 diabetes.

**Methods:** Insufficiently active adults with type 2 diabetes (N=69; 46 females; mean age ± SD: 58±11 years) were randomized to an ES or mobility/stretching comparator group (CON), which involved 4 × 1-min bouts of either vigorous or low intensity exercise, respectively, on ≥5 days/week. The primary outcome was feasibility based on adherence. Secondary outcomes included exercise enjoyment (1-7 scale), rating of perceived exertion (RPE; 0-10 scale), heart rate (HR), hemoglobin A_1c_ (HbA_1c_), blood biomarkers of cardiometabolic health, 30-second sit-to-stand capacity, grip strength, estimated maximal oxygen uptake, and anthropometrics.

**Results:** Weekly adherence (estimated marginal mean [95% confidence interval]: 18 bouts [16 to 21] for both groups; P=0.99) and total enjoyment (ES: 4.5 [4.1 to 4.8] vs CON: 4.3 [4.0 to 4.7]; P=0.64) were high and not different between groups. Despite higher RPE (5.7 [5.4 to 6.1]) and peak HR (73 [70 to 77] % of age-predicted HR maximum) in ES vs CON (2.0 [1.7 to 2.4] and 61 [58 to 64] % of age-predicted HR maximum, respectively) (all P<0.001), there were no between-group differences in the change in any secondary outcome (all P>0.05) except for greater sit-to-stand capacity in ES after training (between-group effect estimate [95% confidence interval]: 1.9 repetitions [0.3 to 3.4]; P=0.02).

**Conclusions/interpretation:** Exercise snacks were feasible to perform in the real-world and improved physical capacity to a greater extent than CON in adults living with type 2 diabetes.

**Trial registration:** ClinicalTrials.gov ID: NCT06407245

**Research in Context:** *What is already known about this subject?:*

- Exercise snacks (≤1-min bouts of vigorous exercise spaced out across the day) are a time-efficient and practical approach to promote vigorous exercise and break up sedentary time.
- Real-world exercise snack interventions appear feasible for middle-aged and older adults.

*What is the key question?:*

- Are 12 weeks of exercise snacks performed in the real-world feasible for insufficiently active adults living with non-insulin treated type 2 diabetes?

*What are the new findings?:*

- Exercise snacks are feasible for those living with type 2 diabetes to perform unsupervised in the real-world based on high adherence, enjoyment, and participant retention rates.
- Exercise snacks improved 30-second sit-to-stand capacity and reduced waist circumference suggesting enhancements in physical capacity and body composition.

*How might this impact on clinical practice in the foreseeable future?:*

- Exercise snacks could be utilized to help individuals living with type 2 diabetes build a routine or habit of incorporating small amounts of physical activity into their daily lives.
- The improved physical capacity observed in the current study could contribute to lower fall risk and greater lower body strength in those with type 2 diabetes as they age.

## Introduction

It is recommended that individuals living with type 2 diabetes accumulate ≥150 minutes of moderate-to-vigorous intensity aerobic physical activity per week [1, 2] given the benefits of habitual exercise training on glucose regulation [3]. Despite this, a recent study showed that only ∼16% of women and ∼26% of men with type 2 diabetes meet these physical activity guidelines and that there is a high prevalence of device-measured sedentary behaviour in this population [2]. Given the risks associated with physical inactivity and prolonged sitting – which include poor glycemic regulation, elevated cardiometabolic risk, and increased mortality - there is a need for innovative, practical strategies to enhance physical activity engagement and reduce sedentary behaviour in those living with type 2 diabetes [1, 4].

Evidence suggests that the intensity of exercise training may be an important contributor to reductions in hemoglobin A_1c_ (HbA_1c_) and improved glycemic regulation in those with type 2 diabetes [5–7]. “Exercise snacks” are defined as short bouts of vigorous exercise (typically ≤1 min) completed periodically throughout the day [8–11]. Exercise snacks are time-efficient, do not require gym access or specialized equipment, and are commonly performed as simple bodyweight exercises enabling their implementation in real-world settings such as the home or workplace [8, 9, 12]. Exercise snacks may represent an accessible, practical, and convenient approach to promote vigorous exercise and break up sedentary time when compared to traditional exercise training modalities [8, 9, 11]. Recent studies have demonstrated the feasibility of exercise snack training programs performed outside of a laboratory setting in middle-aged and older adults [14–17] but the feasibility of exercise snacks in a real-world setting has never been studied in adults living with type 2 diabetes.

The objective of this study was to assess the feasibility and preliminary efficacy of a 12-week remotely-delivered real world exercise snacks (ES) intervention compared to a mobility/stretching exercise comparator (CON) intervention in individuals living with type 2 diabetes using a randomized trial design. Feasibility was the primary outcome assessed through intervention adherence. Exercise enjoyment ratings and ratings of perceived exertion (RPE) were also used to assess intervention feasibility. In anticipation of a future larger scale clinical trial, HbA_1c_ was measured as the main exploratory efficacy outcome along with other secondary markers of exercise intensity, cardiometabolic health, and physical capacity.

## Materials and methods

### Study overview, sample size, and participants

This randomized trial was conducted between April 2024 – August 2025 at McMaster University (Hamilton, Ontario, Canada) and The University of British Columbia (UBC; Kelowna, British Columbia, Canada). This study received approval from the Hamilton Integrated Research Ethics Board (16834) and the UBC Clinical Research Ethics Board (H21-03417) and was pre-registered on ClinicalTrials.gov (NCT06407245). Fig. 1 summarizes the study design. Guidelines for estimating sample size for feasibility and pilot trials are lacking, however, a previous audit of sample sizes in such trials reported medians of n=36 and n=30 per arm, respectively [18]. To complement this, we performed a sample size estimate based on glucose lowering effects in previous literature to detect a within-between interaction for a two-group design with two timpeoints. Lab-based studies involving short exercise bouts to break up sedentary time throughout the day in people with type 2 diabetes have observed large effect sizes (Cohen’s d>0.8) for several glycemic regulation outcomes [19, 20]. Given the real-world nature of our intervention, we selected a more conservative medium effect size (Cohen’s d=0.5, f=0.25). With 90% power, a conservative repeated measures correlation of r=0.3, and two-tailed α-level of 0.05 (G*Power v3.1) indicated 31 participants per group were required. We aimed to recruit 40 participants per group (n=80 total) from the geographical areas around each study site primarily through advertisements by a third-party paid service (Wayturn; Mariefred, Sweden). Participant eligibility criteria are presented in Table 1.

**Figure 1.**
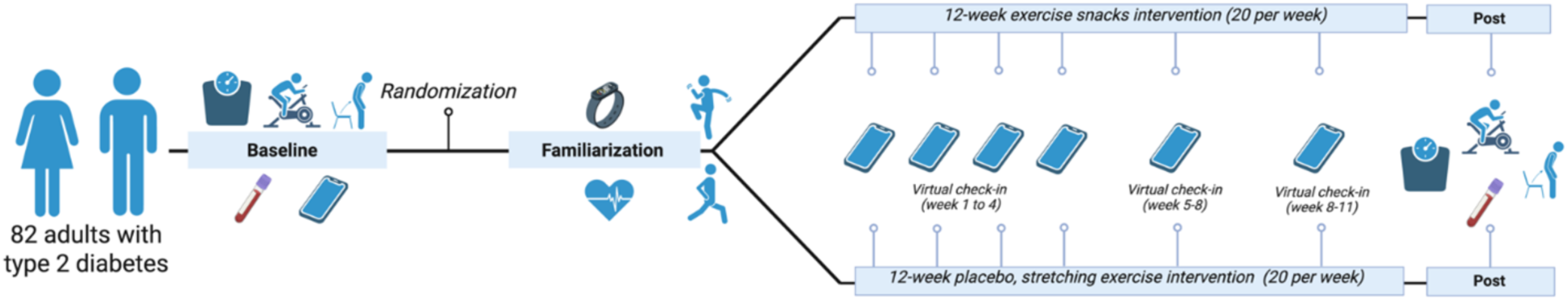
Study design schematic. Baseline and post testing measures included anthropometrics, questionnaires, fasted blood sampling, grip strength, 30-second sit-to-stand, and the YMCA submaximal predictive fitness test. Following these baseline measures, participants were randomized to the exercise snacks or the placebo, exercise stretching, comparator group before completing the exercise familiarization where heart rate was measured using the Polar chest strap at both sites and the Fitbit Charge 6 simultaneously at the McMaster site. During the 12-week intervention, participants received six phone call check-ins. After the 12-week intervention, participants returned to their respective study site for post-testing measurements. This figure was created with BioRender.com.

**Table 1.** Participant inclusion and exclusion criteria.

|  |  |
| --- | --- |
| **Criteria for inclusion** | 30-75 years old, physician-diagnosed type 2 diabetes, performing <150 minutes of moderate-to-vigorous intensity aerobic exercise per week, body mass index (BMI) between 18.5-40 kg/m<sup>2</sup>, no changes to prescribed medications for at least 6 months, able to maintain current medication doses, able to maintain current physical activity patterns, HbA<sub>1c</sub> ≤8.5% (≤69 mmol/mol), access to a computer, tablet, or smartphone, able to travel to a study site for laboratory testing visits, able to read, write, and understand English, anticipated internet access throughout the intervention, and cleared to participate in exercise based on the Canadian Society for Exercise Physiology Get Active Questionnaire [34]. |
| **Criteria for exclusion** | Taking ≥4 glucose-lowering medications, taking insulin, taking beta-blockers, taking ≥3 prescribed medications for the prevention of cardiovascular disease (e.g., statins, antihypertensives), episode of severe hypoglycemia in the past 6 months, cigarette smoker, chronic musculoskeletal condition that would prevent participation in exercise, recent (within the last 2 years) cardiovascular event that prevents participation in exercise, angina upon exertion (self-reported and screened for using survey questions based on those in the Rose Cardiovascular Questionnaire V1 Section A [35]), uncontrolled hypertension or an atypical blood pressure or pulse rate at rest or during exercise as determined by a physician, surgical procedure scheduled during the intervention that would prevent exercise participation, current diagnosis of a cardiac or pulmonary disease that would prevent exercise participation, a psychiatric disorder that could prevent completion of the study procedures or visits, donated >0.5 L of blood within the last 4 weeks, following an extreme diet (e.g., very low carbohydrate/calorie, ketogenic), taking dietary/nutritional supplements that impact glucose control (e.g., exogenous ketones), having diabetic ulcers, peripheral vascular disease, or diabetic neuropathy that would prevent participation in exercise, undergoing dialysis treatment, participating in another clinical trial that would interfere with the study procedures, and pregnant or planning to become pregnant during the intervention. |

### Study protocol

#### Screening

Interested individuals completed a screening call to discuss eligibility criteria review material in the consent form. If deemed eligible based on self-reported responses, an in-person baseline testing visit was scheduled.

#### Baseline testing and exercise familiarization

Individuals arrived after a ≥8-hour fast and provided signed informed consent before baseline measures were collected. Participants were instructed to refrain from taking glucose-lowering medication on the morning of the baseline visit. Height, body mass, and waist circumference were measured first and body mass index (BMI) (kg/m^2^) was calculated. Subsequently, a fasted venous blood sample was obtained via venipuncture and collected into ethylene-diaminetetraacetic acid (EDTA) tubes. HbA_1c_ was measured using a point-of-care multi-assay analyzer (Afinion 2, Abbott Laboratories, Chicago, Illinois, USA) from whole blood. Plasma was obtained by centrifugation at 2500 g for 15 minutes at 4 ° C and stored at −80 ° C prior to batch analyses. Participants were offered a snack supplied by the researcher ≥15 minutes before the next testing procedures. Participants completed an entry survey, which included questions based on demographics, health, and the Godin-Shephard Leisure-Time Physical Activity Questionnaire [21]. Participants then completed a maximal grip strength test involving 2-3 attempts on each hand using a hand dynamometer (Lafayette Instrument Professional Grip Dynamometer, Lafayette, Indiana, USA). Maximum grip strength was the sum of the highest attempt from each hand. Participants then performed a sit-to-stand test which involved repeating as many sit-to-stands as possible within 30 seconds from a chair with no armrests. Participants then completed the YMCA submaximal exercise test [22, 23] on a cycle ergometer (McMaster: Lode Excalibur Sport V 2.0; Lode BV, Groningen, The Netherlands and UBC: SCIFIT ISO1000R; SCIFIT Systems Inc., Tulsa, Oklahoma, USA) to estimate maximal oxygen uptake (VO_2max_) and cardiorespiratory fitness. The test started at 25 watts (W) for 3-min and the heart rate (HR) response to this stage determined the workloads for the subsequent 3-min stages [23]. The aim was for participants to complete two stages that elicit a steady-state HR between 110 bpm and 85% of their age-predicted HR maximum (APHRM; 220-age). HR was measured with a chest strap (Polar, Finland). Participants were then randomized (stratified by study site and sex) to the ES or CON group by a study investigator through the Research Electronic Data Capture (REDCap) system (Vanderbilt University, Nashville, Tennessee, USA). The randomization schedule was created by a third-party researcher and loaded into REDCap in advance. After randomization, participants completed 1-2 familiarization exercise bouts while HR was measured via the Polar chest strap. At the McMaster site, HR was also simultaneously measured with a wrist-worn Fitbit Charge 6 (Fitbit, San Francisco, California, USA). Participants were asked to rate the session RPE (0-10 Session RPE scale [24]) after completing each exercise bout. HR and RPE were averaged if the participant completed two familiarization exercises.

#### Exercise intervention

Participants completed the 12-week remote exercise intervention at home or work with push notifications delivered through a custom smartphone application (Seven Movements, Canmore, Alberta, Canada). The app provided two different instructional exercise videos daily; these were rotated to allow participants to experience a variety of different exercises. Participants pre-selected notification times to complete at least four isolated 1-min exercise bouts per day separated by >1 hour, on at least five days per week. The intervention prescription therefore involved a minimum of 20 weekly 1-min exercise bouts. Participants were encouraged to perform a bout within 30-60 min after major meals when possible. Participants were prompted to complete a post-activity assessment of session RPE [24] and exercise enjoyment (Exercise Enjoyment Scale [25, 26]) on the app following each bout. Participants were given a Fitbit Charge 6 to wear throughout the intervention to measure HR responses to the 1-min exercise bouts. The only difference between the two intervention groups were the types of exercises prescribed. The ES group were assigned higher-intensity bodyweight exercises (e.g., squats, lunges, step-ups, etc.) and this group was referred to as the “high-intensity movement break group” to participants. The ES group was instructed to adjust the pace/effort of the exercises to achieve an RPE of ≥7, which corresponds to “very hard” [24]. The CON group was assigned lower-intensity stretches and mobility movements and was referred to as the “low-intensity movement break group”. A full list of exercises prescribed is listed in Supplementary File 1. Six phone call check-ins were scheduled throughout the intervention: once a week during the first 4 weeks and then once more during both weeks 5-8 and 9-12. The phone calls helped to promote intervention adherence by checking in on exercise progress for the week (participants were asked to self-report the number of bouts performed over the past week) and providing support with any issues encountered with the intervention. For some participants, exercises were removed from the program based on preferences and/or limitations discussed before the start of the intervention or during the phone call check-ins.

#### Post-intervention testing

Twelve weeks after starting the intervention, participants returned to the laboratory for follow-up testing. Baseline testing measures were repeated and participants completed an exit survey, which included questions based on health and the Godin-Shephard Leisure-Time Physical Activity Questionnaire [21].

### Outcomes

#### Intervention feasibility: adherence and enjoyment

App-based adherence during weeks 1-12 was the primary feasibility outcome. Feasibility was defined by the research team *a priori* as >70% of participants completing at least 67% (two-thirds) of the exercise bouts (i.e., >13 weekly bouts) on at least 8 out of 12 weeks. Rates of recruitment, drop-out, retention, exercise enjoyment, and other adherence metrics were used as additional indicators of intervention feasibility. The other markers of adherence included: (1) the total number of bouts completed on the app; (2) the total number of bouts recorded on the Fitbit; (3) the weekly average number of bouts recorded on the Fitbit during weeks 1-12; (4) the weekly average number of self-reported exercise bouts completed on 6 weeks of the intervention (reported during the phone call check-ins). Total app-based adherence during weeks 1-12 was also classified as high (>70% = >168 bouts), moderate (50-69% = 168-120 bouts), or low (<50% = <120 bouts). The total number of app or Fitbit recorded bouts were not included for participants who did not complete the entire intervention period.

#### Exercise intensity

The average RPE was calculated as the mean of all app-recorded RPE ratings during the intervention as an additional feasibility outcome. Second-by-second HR files and activity logs recorded by the Fitbit were downloaded for all participants who registered on Fitabase (n=44) (San Diego, California, USA). Activity logs were cleaned in Python according to the following parameters: manually recorded, 30-180 seconds in duration, recorded between the training start date and post-testing date. The time stamp of each activity enabled identification of peak HR for each activity log with corresponding HR data. The number of Fitbit activity logs was used to calculate Fitbit weekly and total adherence.

#### Cardiorespiratory fitness and leisure score index

VO_2max_ was estimated (see Supplementary File 2) [23] based on a predictive equation using the HR achieved during two stages of the YMCA test where the steady-state HR was between 110 bpm and 85% of APHRM (VO_2max_ ≥110 bpm). Due to the low number of participants who completed a full valid test with two stages in the desired HR range, we explored the estimated VO_2max_ with the same equation method [23] but without the lower HR limit parameter of 110 bpm (VO_2max_ no lower HR limit). We also explored the submaximal HR response for participants who completed the first 25 W workload stage to assess a potential training response to a standardized workload. The Godin-Shephard leisure score index calculation is described in Supplementary File 3. Godin-Shephard responses were removed from analysis as input errors if the number of mild/light reported bouts per week was >60 or if the number of moderate reported bouts per week surpassed eligibility criteria activity level.

#### Plasma analyses

Fasted plasma samples were analyzed for glucose (Human Glucose Assay Kit, Chrystal Chem), insulin (Insulin Elisa Kit, Chrystal Chem), and cytokines (interleukin-6, interleukin-10, tumor necrosis factor-alpha; high-sensitivity VPlex assay, MesoScale Discovery).

### Statistical methods

Baseline characteristics are presented as mean ± standard deviation for continuous variables or N (%) for categorical variables. The preregistered primary outcome of intervention feasibility was assessed descriptively as >70% of participants completing at least 67% of the exercise bouts (i.e., >13 weekly bouts) on at least 8 out of 12 weeks. Adherence and intensity-related outcomes assessed across the intervention period (e.g., mean/total number of bouts, enjoyment, RPE, and HR) or during the exercise familiarization (e.g., RPE and HR) were analyzed using a linear model with a fixed effect for group assignment and accounting for stratified allocation factors (i.e., sex and site). Preliminary efficacy outcomes collected at baseline and follow-up were analyzed using constrained longitudinal data analysis via a linear mixed model with fixed effects for timepoints, the interaction between timepoint and intervention group, stratified allocation factors (i.e., sex and site), and a random effect for participant. Model assumptions of normality and linearity were assessed visually via inspection of diagnostic plots. Models were run using log-transformed outcome variables if departure from model assumptions were observed. Data were analyzed on an intention-to-treat basis, and no statistical imputations were used to replace missing data. Data are presented as model-derived estimated marginal means and within- and between-group effect estimates with corresponding 95% confidence intervals (CI) and P values. Data analyses were conducted in R (version 4.3.2) and statistical significance was established at a two-sided ɑ = 0.05, with no adjustments applied to secondary and exploratory outcomes.

## Results

### Participant recruitment and retention

The participant CONSORT flowchart is displayed in Fig. 2. In total, 358 individuals expressed interest in the study through Wayturn and the other listed modes with N=82 participants recruited (n=42 at McMaster and n=40 at UBC). Baseline measures revealed that seven individuals did not qualify to participate despite self-reporting meeting the eligibility during screening, resulting in 75 participants randomized (ES: n=35 vs CON: n=40). After randomization, six participants were also found to be ineligible based on observations made or information disclosed during baseline testing, resulting in 69 participants (ES: n=31 vs CON: n=38) starting the study intervention and included in the participant baseline characteristics (Table 2). Additional health and demographic information are presented in Supplementary File 4 and data separated by sex are displayed in Supplementary File 5. Of the 69 participants who started the intervention, 63 completed the study and attended follow-up testing, representing a 91% retention rate (63/69).

**Figure 2.**
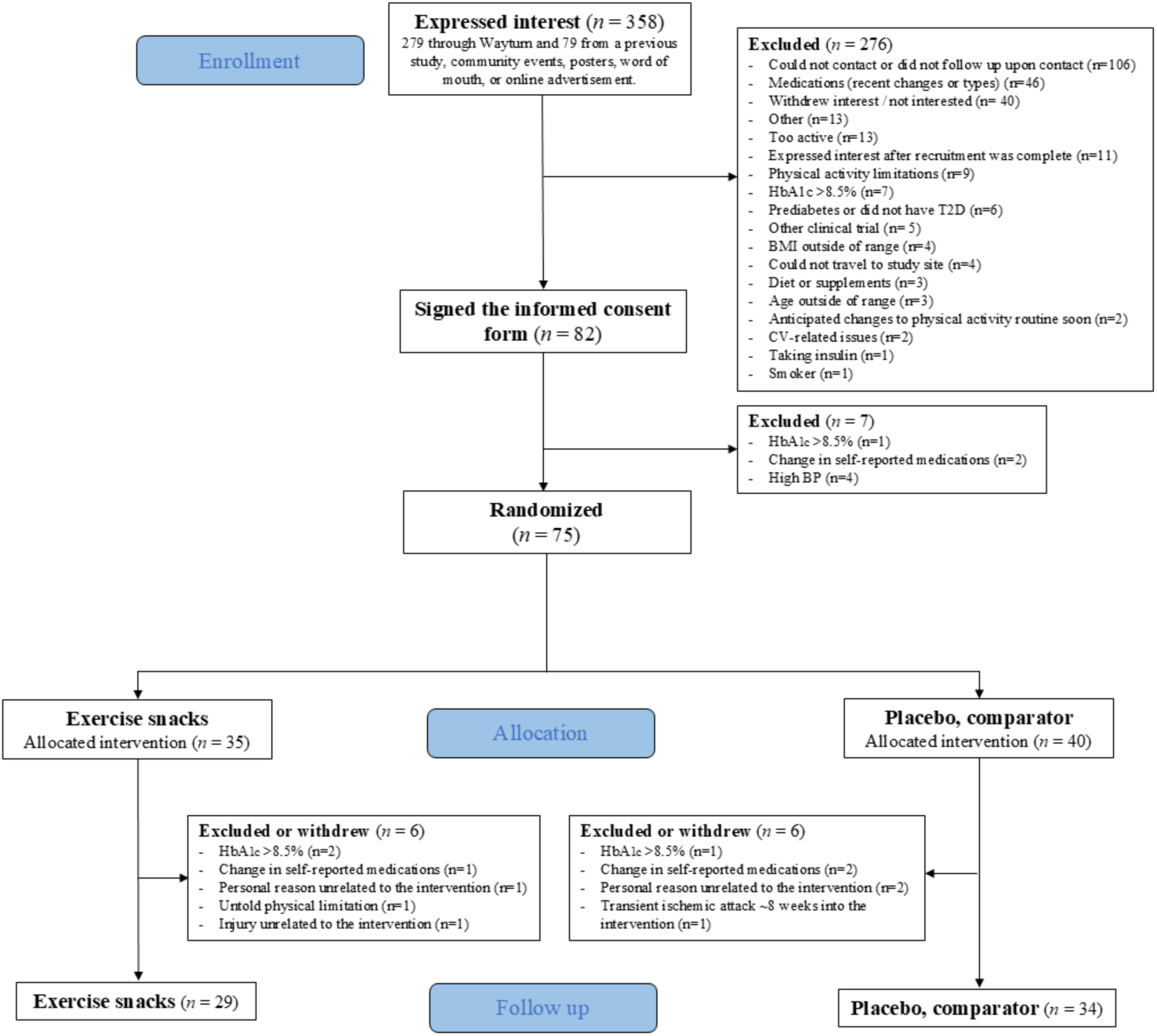
Participant CONSORT flowchart.

**Table 2.** Participant baseline characteristics.

|  | **Total** | **ES** | **CON** |
| --- | --- | --- | --- |
| *N* | 69 | 31 | 38 |
| *Female, N (%)* | 46 (67) | 21 (68) | 25 (66) |
| *Age, yrs* | 58 (11) | 58 (11) | 58 (11) |
| *Anthropometrics* |  |  |  |
| Body mass, kg | 84.7 (16.5) | 89.4 (18.3) | 80.9 (14.1) |
| Body mass index, kg/m<sup>2</sup> | 30.1 (5.4) | 31.4 (5.5) | 28.9 (4.9) |
| Waist circumference, cm | 102 (16) | 105 (19) | 99 (12) |
| *Fitness* |  |  |  |
| VO<sub>2max</sub> ≥110 bpm, mL/kg/min | 25.9 (6.4) | 24.7 (7.9) | 27.2 (4.4) |
| VO<sub>2max</sub> no lower HR limit, mL/kg/min | 23.4 (5.6) | 23.3 (5.8) | 23.5 (5.4) |
| Submaximal HR at 25 W, bpm | 97 (13) | 99 (12) | 96 (13) |
| Grip strength, kg | 74.3 (25.2) | 72.7 (24.0) | 75.4 (26.5) |
| Sit-to-stand 30 sec, repetitions | 13 (4) | 13 (3) | 14 (4) |
| *Metabolic health* |  |  |  |
| HbA<sub>1c</sub>, % | 6.6 (0.8) | 6.6 (0.7) | 6.6 (0.9) |
| Glucose, mmol/L | 6.5 (1.4) | 6.3 (0.9) | 6.6 (1.7) |
| Insulin, mU/L | 10.0 (7.0) | 10.5 (6.9) | 10.0 (8.0) |
| Tumor necrosis factor-alpha, pg/mL | 1.0 (0.3) | 0.9 (0.4) | 1.0 (0.3) |
| Interleukin-6, pg/mL | 0.6 (0.8) | 0.7 (1.1) | 0.6 (0.5) |
| Interleukin-10, pg/mL | 0.2 (0.2) | 0.2 (0.1) | 0.2 (0.3) |
| *Physical activity* |  |  |  |
| Godin-Shephard, leisure score index | 19 (18) | 18 (19) | 20 (17) |

Data presented as mean (SD) unless otherwise specified. ES = exercise snacks group, CON = stretching comparator group; VO<sub>2max</sub> = estimated maximal oxygen uptake; HR = heart rate; W = watts; HbA<sub>1c</sub> = hemoglobin A<sub>1c</sub>. Baseline values were based on n=69 unless otherwise noted. Waist circumference: n=66; VO<sub>2max</sub> ≥110 bpm: n=19; VO<sub>2max</sub> no lower HR limit: n=44; submaximal HR at 25 W: n=61; grip strength: n=53; sit-to-stand: n=68; glucose, insulin, tumor necrosis factor-alpha, interleukin-6, and interleukin-10: n=62; Godin-Shephard: n=68.

### Feasibility: adherence and enjoyment

App-based adherence showed that 77% (48/62) of participants completed >13 exercise bouts (i.e., >67% of the weekly exercise prescription) on ≥8 of the 12 intervention weeks. Weekly app-based adherence is displayed in Fig. 3. The mean weekly number of exercise bouts recorded on the app was not different between ES and CON (estimated marginal means [95% CI]: 18 bouts [16 to 21] in both groups; P=0.99; n=64). Total exercise bouts completed on the app were not different between ES (230 bouts [199 to 262]) and CON (231 bouts [202 to 260]) (P=0.96; n=62). Based on the total bouts recorded on the app between weeks 1-12, 49 participants (∼79%) fell into the high adherence category (>70% = >168 bouts), 8 participants (∼13%) had moderate adherence (50-69% = 168-120 bouts), and 5 participants (∼8%) had low adherence (<50% = <120 bouts). The total (ES: 190 bouts [145 to 235] vs CON: 165 bouts [128 to 203]; P=0.36; n=43) and weekly (ES: 16 bouts [12 to 20] vs CON: 13 bouts [10 to 16]; P=0.25; n=44) number of exercise bouts recorded on the Fitbit were also not different between groups. The weekly average number of exercise bouts self-reported during the phone call check-ins was also not different between groups (ES: 20 bouts [19 to 22] vs CON: 20 bouts [19 to 21]; P=0.86; n=65). Exercise enjoyment, on a scale of 1-7, were not different between groups (ES: 4.5 [4.1 to 4.8] vs CON: 4.3 [4.0 to 4.7]; P=0.64; n=64).

**Figure 3.**
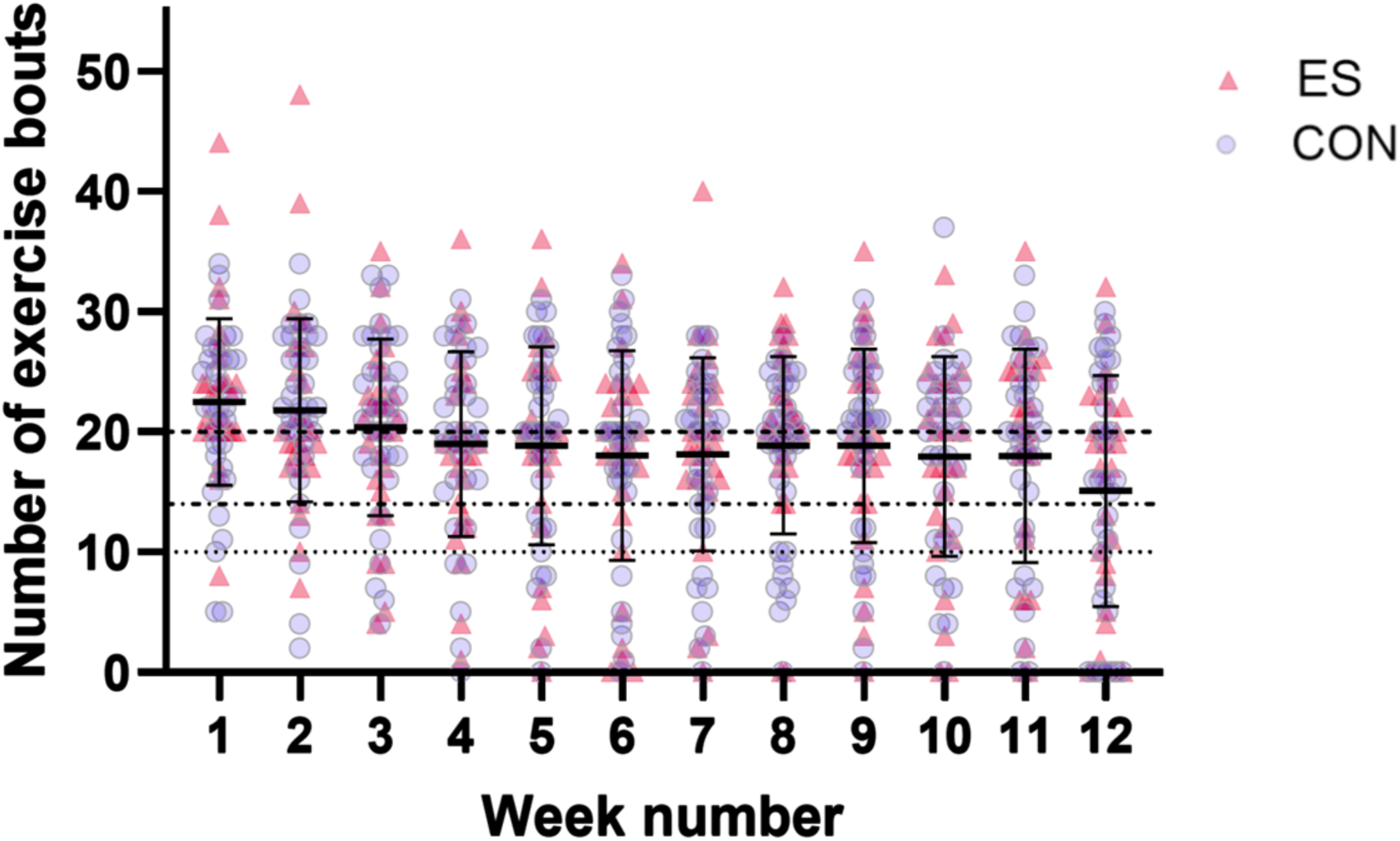
Weekly app-based exercise adherence. Line and error bars shown for each week display the mean ± standard deviation. The three horizontal dashed/dotted lines across the graph represent (1) the prescribed weekly number of exercise bouts (y=20), (2) the breakpoint between high (>70%) and moderate (50-69%) adherence (y=14), and (3) the breakpoint between moderate and low (<50%) adherence (y=10). ES = exercise snacks group, CON = stretching comparator group. Week 1-3 are based on n=64, week 4-8 are based on n=63, and week 9-12 are based on n=62.

### Exercise intensity: heart rate and RPE

During familiarization, RPE was 6.3 (5.8 to 6.7) in the ES group vs 1.6 (1.0 to 2.1) in the CON group with a between-group difference of 4.7 (4.0 to 5.4) (P<0.001; n=45). Peak HR measured with the Polar chest strap during the familiarization was 133 bpm (127 to 139) or 81% (78 to 84) of APHRM in the ES group compared to 99 bpm (94 to 104) or 62% (59 to 64) of APHRM in the CON group (P<0.001; n=63). Peak HR measured with the Fitbit during the exercise familiarization was 132 bpm (125 to 140) or 80% (76 to 84) of APHRM in the ES group compared to 94 bpm (88 to 101) or 57% (53 to 61) of APHRM in the CON group (P<0.001; n=32).

During the intervention, RPE was higher in the ES group (5.7 [5.4 to 6.1]) compared to the CON group (2.0 [1.7 to 2.4]) with a between-group difference of 3.7 (3.2 to 4.2) (P<0.001; n=64). Peak HR recorded with the Fitbit during the intervention was higher in the ES group (121 bpm [113 to 128]) vs the CON group (97 bpm [91 to 104]) (P<0.001; n=41). This corresponded to 73% (70 to 77) of APHRM in the ES group vs 61% (58 to 64) in the CON group (P<0.001; n=41). RPE and HR during weeks 1-12 of the intervention are displayed in Fig.4.

**Figure 4.**
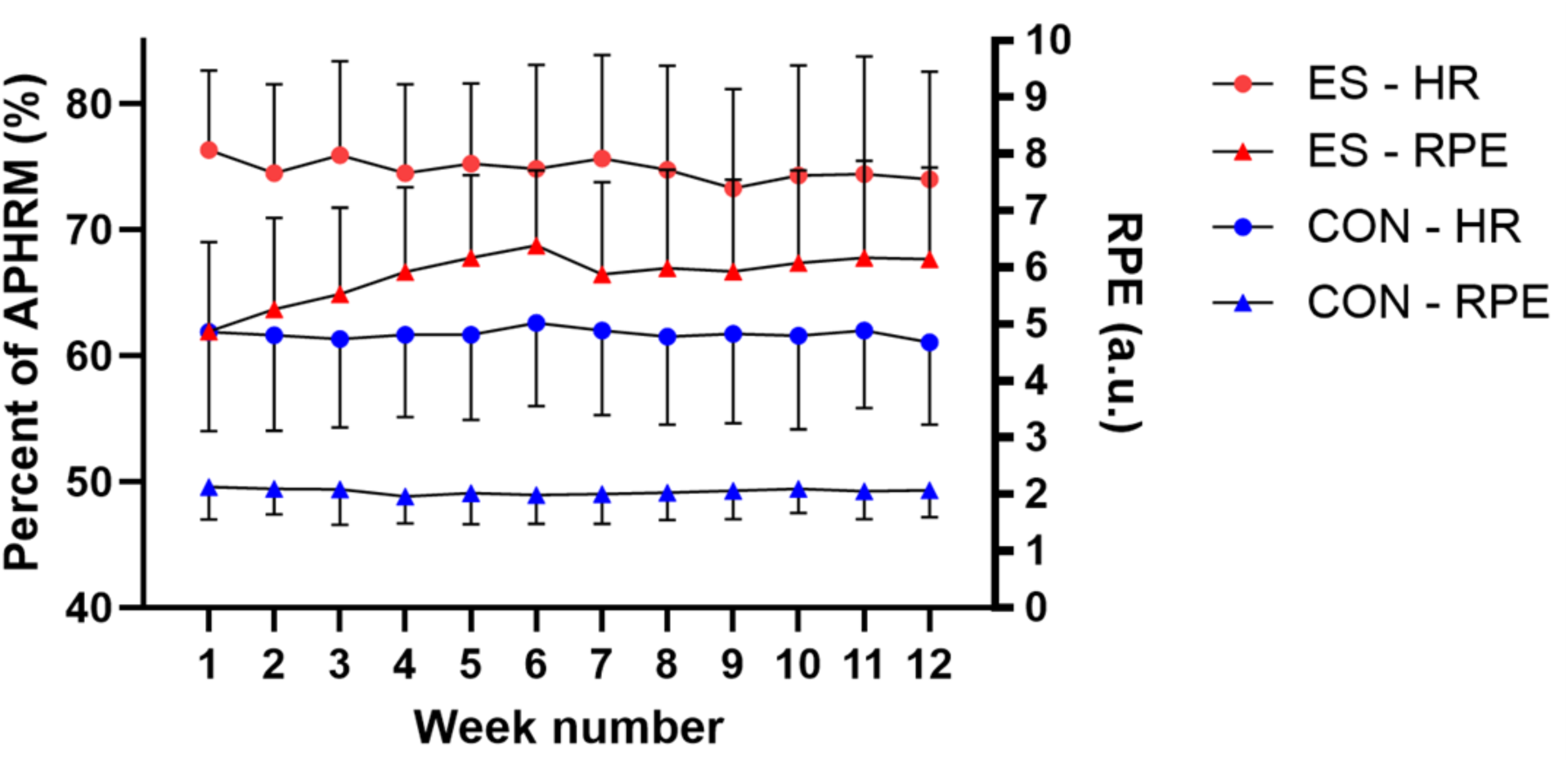
Weekly exercise intensity during the 12-week intervention. Peak heart rate (HR) responses recorded with the Fitbit for exercise bouts shown as percentages of age-predicted HR maximum (APHRM) (left y-axis) and session rating of perceived exertion (RPE) reported on the app after completing an exercise bout (right y-axis). ES = exercise snacks group, CON = stretching comparator group. ES RPE are based on n=25-29, ES HR are based on n=14-18, CON RPE are based on n=27-35, and CON HR are based on n=18-22 depending on the number of individuals who recorded RPE and HR during each week of the intervention.

### Efficacy outcome measures

Within- and between-group effect estimates and 95% CI for all clinical and physical outcomes are presented in Table 3. There were no between-group differences in the change in any clinical efficacy measure including HbA_1c_, fasting glucose, fasting insulin, plasma cytokines, body mass, BMI, or waist circumference in response to the intervention (P>0.05). There was an increase in 30-second sit-to-stand capacity in the ES and CON group (within-group effect estimates [95% CI]: 4.4 repetitions [3.2 to 5.7] vs 2.6 repetitions [1.5 to 3.6], respectively; both P<0.001), resulting in a between-group difference indicating a greater increase in the ES group (1.9 repetitions [0.3 to 3.4]; P=0.02). There was a within-group reduction in waist circumference in the ES group from baseline to follow-up (−1.6 cm [−3.2 to −0.1]; P=0.04). There were no between-group differences observed in the change in the other physical parameters measured including grip strength, estimated VO_2max_, or HR responses to submaximal exercise (P>0.05). There were also within-group changes observed for the Godin-Shephard leisure score index in both ES (14 [8 to 20]; P<0.001) and CON (7 [2 to 13]; P=0.01), with no between-group differences (P>0.05).

**Table 3.** Effect estimates for changes in outcomes measures.

|  | Change in ES | Change in CON | Difference Between Groups |
| --- | --- | --- | --- |
| *Anthropometrics* |  |  |  |
| Body mass, kg | -0.7 (-1.5 to 0.1), P=0.09 | -0.7 (-1.4 to 0.0), P=0.06 | 0.0 (-1.0 to 1.1), P=0.96 |
| Body mass index, kg/m<sup>2</sup> | -0.2 (-0.5 to 0.1), P=0.15 | -0.2 (-0.5 to 0.0), P=0.09 | 0.0 (-0.4 to 0.4), P=0.91 |
| Waist circumference, cm | -1.6 (-3.2 to -0.1), P=0.04 | -0.2 (-1.6 to 1.2), P=0.75 | -1.4 (-3.5 to 0.7), P=0.18 |
| *Fitness* |  |  |  |
| VO<sub>2max</sub> ≥110 bpm, mL/kg/min | -1.0 (-5.0 to 3.0), P=0.60 | -1.5 (-5.8 to 2.8), P=0.47 | 0.5 (-5.1 to 6.1), P=0.85 |
| VO<sub>2max</sub> no lower HR limit, mL/kg/min | 0.1 (-1.7 to 1.9), P=0.91 | -0.9 (-3.1 to 1.3), P=0.43 | 1.0 (-1.7 to 3.7), P=0.47 |
| Submaximal HR at 25 W, bpm | -3.2 (-6.9 to 0.6), P=0.10 | -3.1 (-6.8 to 0.5), P=0.09 | 0.0 (-5.0 to 4.9), P=0.99 |
| Grip strength, kg | 2.0 (-0.6 to 4.7), P=0.13 | 0.3 (-2.1 to 2.6), P=0.83 | 1.8 (-1.7 to 5.3), P=0.31 |
| Sit-to-stand 30 sec, repetitions | 4.4 (3.2 to 5.7), P<0.001 | 2.6 (1.5 to 3.6), P<0.001 | 1.9 (0.3 to 3.4), P=0.02 |
| *Metabolic health* |  |  |  |
| HbA<sub>1c</sub>, % | -0.1 (-0.3 to 0.1), P=0.23 | 0.0 (-0.2 to 0.1), P=0.62 | -0.1 (-0.3 to 0.2), P=0.57 |
| Glucose, mmol/L | -0.1 (-0.6 to 0.3), P=0.62 | 0.1 (-0.3 to 0.5), P=0.69 | -0.2 (-0.8 to 0.4), P=0.51 |
| Insulin, mU/L<sup>1</sup> | 5 (-16 to 31), P=0.68 | 1 (-17 to 24), P=0.90 | 3 (-23 to 38), P=0.82 |
| Tumor necrosis factor-alpha, pg/mL<sup>1</sup> | 0 (-7 to 8), P=0.94 | -5 (-11 to 1), P=0.09 | 6 (-3 to 16), P=0.22 |
| Interleukin-6, pg/mL<sup>1</sup> | 4 (-16 to 29), P=0.70 | 2 (-17 to 24), P=0.87 | 3 (-22 to 35), P=0.85 |
| Interleukin-10, pg/mL<sup>1</sup> | -4 (-23 to 20), P=0.69 | -17 (-33 to 2), P=0.08 | 15 (-13 to 53), P=0.32 |
| *Physical activity* |  |  |  |
| Godin-Shephard, leisure score index | 14 (8 to 20), P<0.001 | 7 (2 to 13), P=0.01 | 7 (-1 to 14), P=0.09 |

All data are presented as (within- or between-group) effect estimates (95% confidence interval) with corresponding P values. Effect estimates are based on intention-to-treat analyses. Data analyzed via constrained longitudinal data analysis using a linear mixed model with fixed effects for timepoints, the interaction between timepoint and intervention group, stratified allocation factors (sex and site), and a random effect for participant. ES = exercise snacks group, CON = stretching comparator group; VO<sub>2max</sub> = estimated maximal oxygen uptake; HR = heart rate; W = watts; HbA<sub>1c</sub> = hemoglobin A<sub>1c</sub>. Post testing values were based on n=63 unless otherwise noted; n=1 from the ES group experienced a high blood pressure episode at home near the end of the intervention which impacted the collection of certain post testing measurements for this participant. Waist circumference: n=54; VO<sub>2max</sub> ≥110bpm: n=15; VO<sub>2max</sub> no lower HR limit: n=35 (n=1 removed from the dataset because calculated VO<sub>2max</sub> was outside of reasonable physiologic range); submaximal HR at 25 W: n=58; grip strength: n=46; sit-to-stand: n=61; HbA<sub>1c</sub>: n=60; glucose, insulin, tumor necrosis factor-alpha, interleukin-6, and interleukin-10: n=58;
<sup>1</sup> Log-transformed, interpret as percent change.

## Discussion

The primary finding of the present trial was that 12 weeks of 4 × 1-min daily movement breaks (exercise snacks and stretching exercises) delivered via app and performed unsupervised in a real-world setting, were feasible to complete for insufficiently active adults living with type 2 diabetes. We observed that 77% of participants performed >13 exercise bouts on ≥8 of the 12 intervention weeks, which exceeded our pre-defined threshold of >70% adherence. We also found a greater increase in 30-second sit-to-stand capacity in the ES group compared to the CON group and a within-group reduction in waist circumference in the ES group after training, suggesting improved physical capacity and body composition with vigorous exercise snacks. However, there were no apparent between-group differences in other clinical efficacy markers assessed, including those related to glycemic regulation, in this cohort of participants with well-controlled glycemia at baseline.

To our knowledge, this is the first trial to demonstrate the feasibility of a real-world exercise snacks intervention in those living with type 2 diabetes, which was determined through the adherence, enjoyment, and retention rates (>90%). Total app-based adherence revealed that 79% of participants were high adherers to the intervention, completing >168 exercise bouts across 12 weeks. Weekly app-based adherence and self-reported adherence were high (between 18-20 bouts per week and not different between groups), which closely aligns with the exercise prescription of 20 weekly bouts. Consistent with our findings, previous home-based exercise snack intervention studies in non-type 2 diabetes participants support high exercise adherence and the feasibility of this approach [14–17]. In the current study, exercise enjoyment were between 4-5 out of 7 representing “moderate” to “quite a bit” on the Exercise Enjoyment Scale [25, 26] which coincides with those previously reported in exercise snacks studies performed in lab [27] and real-world [17] settings. Furthermore, participants performed the exercise snacks at a greater exercise intensity compared to previous real-world exercise snack interventions [14, 17]. RPE was previously ≤4/10 [14, 17] compared to ∼6/10 in the current study. The RPE in the present study aligns with the RPE reported in previous lab-based exercise snack interventions, which were between ∼5-7 on a 0-10 RPE scale [27–29]. Although not captured in all participants, the 1-min exercise snacks were associated with a peak HR response of ∼73% of APHRM, suggesting that participants approached common vigorous-intensity thresholds of >76% HR maximum [30]. Together, these findings support the feasibility of a remotely-delivered app-based exercise snacks intervention to increase engagement in short bouts of vigorous exercise in people living with type 2 diabetes.

We did not observe any between-group differences in the change in selected clinical outcome measures of HbA_1c_, blood markers of cardiometabolic health, body mass, BMI, or waist circumference. Although a previous highly-controlled lab-based study found that hourly stair-climbing exercise snacks could improve indices of insulin sensitivity across a 9-hour experimental condition in people with overweight or obesity [31], markers of metabolic health measured before and after the 12-week intervention did not appear to be altered in the present study. It is possible that acute metabolic changes to exercise snacks do not translate to chronic adaptations or that the small dose of prescribed exercise in the current study is not sufficient to elicit improvements in glycemic regulation. Our sample had well-controlled glucose at baseline (mean HbA_1c_ of ∼6.6% and mean fasting glucose of 6.5 mmol/L) which may have limited the ability to see improvements. Similarly, we did not observe between-group differences in the change in most of the physical outcome measures (predicted VO_2max_, submaximal exercise HR responses, and grip strength). We did, however, see a greater improvement in 30-second sit-to-stand capacity in the ES group after the intervention, which is a key indicator of lower body strength and fall risk in older adults [32, 33]. The lack of change in the other markers may be attributed to the prescribed exercise dose (duration and/or intensity) or the total intervention length. It is also possible that sit-to-stand capacity is more specific and amenable to improvements in response to the bodyweight exercise snacks prescribed in the current study.

Strengths of this study include the real-world nature of the intervention, the autonomy given to participants to select between two daily exercise choices, the use of push notifications and phone call check-ins to promote adherence, and the implementation of real-time objective monitoring of the exercise snacks using the Fitbit Charge 6. This study was not without limitations. It is possible that the different exercise adherence measurement techniques used may not have fully captured all the exercise bouts completed by the participants. It also appears that despite high adherence and enjoyment, participants may have had difficulty performing exercise snacks at a vigorous enough intensity when unsupervised in the real world, at least based on Fitbit HR responses. The peak HR response during the baseline exercise familiarization in the lab was ∼80% of APHRM compared to ∼73% during the intervention in the ES group. We also found that the use of the submaximal YMCA predictive fitness test for estimating VO_2max_ performed poorly within this study population, with only ∼30% of participants completing a valid test according to the established protocol [22, 23]. Future work should investigate more suitable fitness testing options for this population.

In conclusion, twelve weeks of exercise snacks are feasible to perform in a real-world setting and can improve sit-to-stand capacity in insufficiently active individuals living with type 2 diabetes. Exercise snacks may represent an intervention to increase engagement in vigorous activity and break up sedentary time for those with type 2 diabetes, but future work appears needed to optimize dosing for improving other physical or clinical measures, including HbA_1c_, in those with already well-controlled glycemia. Continued research should investigate different variations of real-world exercise snack interventions to determine if altering the exercise prescription can produce clinically meaningful improvements in health-related outcomes.

## Data availability

Upon reasonable request to the corresponding author, data can be made available.

## Authors’ relationships and activities

MJG is an advisor to and holds equity in Longevity League Ltd., a US-based company whose services in part relate to exercise. JPL is Chief Scientific Officer of the Institute for Personalized Therapeutic Nutrition (a Canadian registered charity) and holds founders shares in Metabolic Insights Inc., a company developing non-invasive metabolic monitoring devices.

## Contribution statement

FJB contributed to the study design, performed data collection, data analysis, interpreted the results, and drafted the manuscript. AMC contributed to the study design, data collection, data analysis, interpreting the results, and drafting the manuscript. RS contributed to data collection, data analysis, and reviewing the manuscript. KF performed data analysis and data interpretation and helped with drafting the manuscript. NW contributed to data collection and reviewed the manuscript. HI contributed to designing the study and reviewed the manuscript. DR participated in data collection and reviewed the manuscript. SM contributed to data analysis, interpreting the results, and reviewed the manuscript. KM, JS, and MR contributed to the study design and reviewed the manuscript. MJ contributed to the study design, including components/elements related to behavioural change, and reviewed the manuscript. MJG contributed to designing the study, data collection, interpreting the results, and drafting the manuscript. JPL contributed to designing the study, data collection, interpreting the results, and drafting the manuscript.

## Abbreviations

APHRM: age-predicted heart rate maximum
CI: confidence interval
CON: stretching comparator group
ES: exercise snacks group
HR: heart rate
REDCap: Research Electronic Data Capture
RPE: rating of perceived exertion
SD: standard deviation
VO_2max_: estimated maximal oxygen uptake
UBC: The University of British Columbia – Okanagan
W: watts

