## Supplementary material for "Exercise snacks are feasible to perform in the real world and improve physical capacity for adults living with non-insulin treated type 2 diabetes: a randomized trial"

Supplementary File 1: List of exercises prescribed in the exercise snacks (ES) and placebo, stretching exercise comparator (CON) group.

| **ES** | **CON** |
|---|---|
| March in Place | Hands On Chair Sit Back |
| Bodyweight Squat | Chest Stretch |
| Step Ups | Side Lying T Spine Stretch |
| Alternating Forward Lunge | Single Seated Arm and Neck Stretch |
| Change of Direction Run | Rotational Stretch at Wall |
| Reverse Lunge Arm Swing | Standing Type 1 Thoracic Spine Reaches |
| Sit to Stand | Staggered Stance Arms Overhead Look to sky |
| Lateral Squat Walk | Ankle Forward Reach "Pet the Bug" |
| Lateral Shuffle | Single Leg Balance at Wall Rotational Knee Driver |
| Alternating Lateral Lunge | Half Kneeling Hip Mobilizer |
| Alternating Transverse Lunge | Prone Pigeon mobilizer |
| Box Run | Seated Butterfly Stretch |
| Carioca | Supine Eagle |
| Alternating Side Lunge with Twist | Supine Hamstring Stretch |

Supplementary File 2: Equations used to predict $VO_{2max}$ from leg ergometry[1]

$VO_{2max}$ (mL/kg/min) = $SM_2 + b * (\text{age predicted } HR_{max} - HR_2)$

- $b = (SM_2 - SM_1) / (HR_2 - HR_1)$

$HR_1$ and $HR_2$ = final heart rate at each submaximal workload (bpm)

$SM_1$ and $SM_2$ = gross $VO_2$ at each submaximal workload (mL/kg/min)

$= [(W / M) * 1.8] + 3.5$

- W = work rate (kgm/min)
- M = body mass (kg)

Supplementary File 3: Godin-Shephard leisure score index.

Participants were asked the following question on the entry and exit survey based on the Godin-Shephard Leisure-Time Physical Activity Questionnaire [1]:

"During a typical 7-day period (a week), how many times on the average do you do the following kinds of exercise for more than 15 minutes during your free time (write on each line the appropriate number)?"

Participants were asked to self-report the number of weekly bouts of mild/light, moderate, and strenuous exercise lasting more than 15 minutes.

The Godin-Shephard leisure score index was calculated using the following equation:

Leisure score index = (number of mild/light bouts * 3) + (number of moderate bouts * 5) + (number of strenuous bouts * 9)

Supplementary File 4: Participant health and demographic information.

| Descriptor | Sub-descriptor | N (responses) | % |
|---|---|---|---|
| Sex | | 69 | |
| | Male | 23 | 33 |
| | Female | 46 | 67 |
| Menopause status | | 45 | |
| | Post menopause | 23 | 51 |
| | Pre menopause | 12 | 27 |
| | Has had a hysterectomy | 7 | 16 |
| | Other | 3 | 7 |
| Ethnicity or cultural background | | 71* | |
| | White | 51 | 72 |
| | South Asian | 8 | 11 |
| | Other | 3 | 4 |
| | Filipino | 2 | 3 |
| | Latin American | 2 | 3 |
| | Native/Aboriginal | 2 | 3 |
| | Arab | 1 | 1 |
| | Black | 1 | 1 |
| | Chinese | 1 | 1 |
| Time since T2D diagnosis | | 69 | |
| | Less than 1 year | 7 | 10 |
| | 1-2 years | 16 | 23 |
| | 3-5 years | 13 | 19 |
| | 6-10 years | 10 | 14 |
| | More than 10 years | 23 | 33 |
| Employment status | | 69 | |
| | Employed full-time | 34 | 49 |
| | Employed part-time | 6 | 9 |
| | Unemployed | 4 | 6 |
| | Retired | 25 | 36 |
| | Student | 0 | 0 |
| | Unable to work | 0 | 0 |
| Highest level of education | | 69 | |
| | Less than high school | 1 | 1 |

| | | | |
|---|---|---|---|
| | High school diploma or equivalent | 3 | 4 |
| | Some college, no degree | 14 | 20 |
| | Associate degree | 9 | 13 |
| | Bachelor's degree | 23 | 33 |
| | Master's degree | 17 | 25 |
| | Professional degree or Doctorate | 2 | 3 |
| Medication Information | | 69 | |
| | Number of participants taking zero medications | 7 | 10 |
| | Number of participants taking medications | 62 | 90 |
| Medication Type | Number of participants taking each type of medication | | |
| Metformin | 41 | | |
| GLP-1RA | 17 | | |
| Other glucose lowering | 24 | | |
| Antihypertensive | 29 | | |
| Lipid-lowering agent | 37 | | |
| Other | 37 | | |

T2D = type 2 diabetes, GLP-1 = glucagon-like peptide-1 receptor agonist.

*Based on n=69 but participants could select multiple applicable answers.

Supplementary File 5: Data separated by sex.

**Participant baseline characteristics.**

| | **Male** | **Female** |
|---|---|---|
| *N* | 23 | 46 |
| *Age, yrs* | 57.7 (9.6) | 57.9 (11.1) |
| *Anthropometrics* | | |
| Body mass, kg | 91.8 (16.4) | 81.2 (15.6) |
| Body mass index, kg/m$^2$ | 29.4 (4.6) | 30.4 (5.6) |
| Waist circumference, cm | 103.7 (14.7) | 102.5 (13.9) |
| *Fitness* | | |
| $VO_{2max}$ ≥110 bpm, mL/kg/min | 28.4 (7.5) | 23.0 (3.6) |
| $VO_{2max}$ no lower HR limit, mL/kg/min | 26.4 (6.4) | 21.5 (4.1) |
| Submaximal HR at 25 W, bpm | 91 (11) | 101 (12) |
| Grip strength, kg | 103.6 (13.4) | 59.2 (14.0) |
| Sit-to-stand 30 sec, repetitions | 14 (3) | 13 (4) |
| *Metabolic health* | | |
| $HbA_{1c}$, % | 6.8 (0.6) | 6.5 (0.8) |
| Glucose, mmol/L | 6.6 (1.5) | 6.4 (1.4) |
| Insulin, mU/L | 7.9 (4.0) | 11.0 (8.4) |
| Tumor necrosis factor-alpha, pg/mL | 0.9 (0.2) | 1.0 (0.4) |
| Interleukin-6, pg/mL | 0.7 (1.3) | 0.6 (0.5) |
| Interleukin-10, pg/mL | 0.2 (0.1) | 0.2 (0.3) |

Data presented as mean (SD) unless otherwise specified. $VO_{2max}$ = estimated maximal oxygen uptake; HR = heart rate; W = watts; $HbA_{1c}$ = hemoglobin $A_{1c}$. Baseline values were based on n=69 unless otherwise noted. Waist circumference: n=66; $VO_{2max}$ ≥110 bpm: n=19; $VO_{2max}$ no lower HR limit: n=44; submaximal HR at 25 W: n=61; grip strength: n=53; sit-to-stand: n=68; glucose, insulin, tumor necrosis factor-alpha, interleukin-6, and interleukin-10: n=62.

**Participant post-testing characteristics.**

| | **Male** | **Female** |
|---|---|---|
| *N* | 21 | 42 |
| *Anthropometrics* | | |
| Body mass, kg | 90.5 (16.2) | 80.0 (15.6) |
| Body mass index, $kg/m^2$ | 29.1 (4.6) | 29.9 (5.7) |
| Waist circumference, cm | 103.5 (13.8) | 99.8 (13.7) |
| *Fitness* | | |
| $VO_{2max}$ ≥110 bpm, mL/kg/min | 25.4 (6.7) | 21.7 (8.1) |
| $VO_{2max}$ no lower HR limit, mL/kg/min | 24.9 (5.8) | 20.7 (5.9) |
| Submaximal HR at 25 W, bpm | 88 (15) | 97 (13) |
| Grip strength, kg | 102.6 (16.0) | 61.8 (13.0) |
| Sit-to-stand 30 sec, repetitions | 18 (6) | 16 (5) |
| *Metabolic health* | | |
| $HbA_{1c}$, % | 6.6 (0.6) | 6.5 (0.9) |
| Glucose, mmol/L | 6.4 (1.2) | 6.5 (2.1) |
| Insulin, mU/L | 14.7 (19.2) | 10.5 (7.7) |
| Tumor necrosis factor-alpha, pg/mL | 1.0 (0.2) | 0.9 (0.4) |
| Interleukin-6, pg/mL | 0.6 (0.6) | 0.7 (0.7) |
| Interleukin-10, pg/mL | 0.2 (0.2) | 0.2 (0.3) |

Data presented as mean (SD) unless otherwise specified. $VO_{2max}$ = estimated maximal oxygen uptake; HR = heart rate; W = watts; $HbA_{1c}$ = hemoglobin $A_{1c}$. Post testing values were based on n=63 unless otherwise noted. Waist circumference: n=54; $VO_{2max}$ ≥110bpm: n=15; $VO_{2max}$ no lower HR limit: n=35 (n=1 removed from the dataset because calculated $VO_{2max}$ was outside of reasonable physiologic range); submaximal HR at 25 W: n=58; grip strength: n=46; sit-to-stand: n=61; $HbA_{1c}$: n=60; glucose, insulin, tumor necrosis factor-alpha, interleukin-6, and interleukin-10: n=58
